# More efficient use of colonoscopy-based colorectal cancer screening by low-barrier, low-threshold pretesting

**DOI:** 10.1101/2023.08.22.23294401

**Authors:** Thomas Heisser, (Postdoctoral Researcher), Rafael Cardoso, (Postdoctoral Researcher), Tobias Niedermaier, (Postdoctoral Researcher), Michael Hoffmeister, (Professor of Epidemiology), Hermann Brenner, (Professor of Epidemiology)

## Abstract

**Background:** ‘Gateopener’ colonoscopy-based screening is an innovative concept to better target colonoscopy to those most likely to benefit. It combines invitations to screening colonoscopy with the offer of pretesting with a single ‘gateopener’ fecal immunochemical test (FIT) which is applied with a lower positivity threshold than in conventional screening. We explored optimized use of this approach for reducing CRC incidence and mortality.

**Methods and Findings:** Using COSIMO, a validated Markov-based simulation tool, we compared outcomes of gateopener screening to those of conventional FIT- or colonoscopy-based screening strategies. Gateopener screening was modelled using SENTiFIT-FOB Gold (Sentinel Diagnostics) as exemplary ‘gateopener’ FIT. We assessed various low hemoglobin cut-offs (10,8,6,4, and 3 µg/g feces). We found that gateopener screening at cut-offs of 6, 4 or 3 µg/g outperformed conventional screening colonoscopy in terms of CRC incidence reduction, with 16-25%, 50-57% and 66-72% more prevented cases, respectively, after ten years. All gateopener scenarios significantly increased prevented deaths, at low cut-offs more than doubling the numbers achieved by conventional screening colonoscopy. Compared to biennial FIT, gateopener screening prevented 7-163% more CRC cases, with lower cut-offs associated with higher gains, and prevented approximately equal to significantly higher (12-21%) numbers of CRC deaths. Cut-offs of 10 and 8 µg/g required fewer colonoscopies per prevented case and death.

**Conclusions:** Gateopener screening outperforms conventional CRC screening by offering considerably stronger reduction of CRC incidence and mortality rates as well as considerably increased screening effectiveness. The feasibility of the concept should be assessed by a pilot study in real-life practice.

**Author Summary:** *Why was this study done?:*

✓ Screening colonoscopy is used inefficiently as most individuals have no benefit from this invasive procedure.
✓ Efficiency could be enhanced by pre-selecting those most likely to benefit, e.g., by use of a single low-threshold fecal immunochemical test (‘gateopener’ FIT).
✓ Such pre-test may lower the barrier against using screening colonoscopy and may better target use of colonoscopy.

*What Did the Researchers Do and Find?:*

✓ A simulation study comparing application of the gateopener approach to conventional colonoscopy or FIT-based screening approaches demonstrated higher proportions of prevented colorectal cancer cases or deaths and/or a more effective use of colonoscopies as compared to conventional screening.

*What Do These Findings Mean?:*

✓ Inviting subjects to undergo pre-testing with low-threshold FITs could markedly improve outcomes and efficiency of colonoscopy-based colorectal cancer screening.

**Lay summary:** This simulation study demonstrates that introducing a simple, highly sensitive pre-test (e.g., an adjusted modern stool test) to identify individuals most likely to benefit from screening colonoscopy could markedly improve the benefits of colorectal cancer screening.

## Introduction

Screening colonoscopy is the gold standard screening test for colorectal cancer (CRC) as it allows for detection and removal of precursor lesions at high sensitivity directly upon examination.[1] However, most screenees have no real benefit from this invasive procedure as they do not have CRC or any such precursor lesions.[2,3] Also, screening colonoscopy programs often fail to achieve adequate uptake,[4] which implies that many of those at high CRC risk are still being missed for not using screening in the first place. Substantially higher uptake could be achieved by non-invasive tests, such as fecal immunochemical tests (FITs),[5] and further enhanced by optimized screening program design and invitation approaches.[6] However, FITs are expected to mainly contribute to reducing CRC mortality but less so to reducing CRC incidence, as sensitivity is high for cancers but suboptimal for precursor lesions.[7,8]

We recently proposed a ‘gateopener’ approach suited to address these limiting factors in an innovative fashion, with great potential to enhance the effectiveness of CRC screening.[9] Gateopener screening involves pre-selecting subjects most likely to benefit, as only individuals with a positive pre-test, such as a low threshold ‘gateopener’ FIT with low barriers to access (e.g., by directly mailing test kits) would be invited to proceed to screening colonoscopy. As illustrated by a numerical example in **Figure 1**, this approach could lead to more targeted use of the same number of colonoscopies compared to conventional untargeted screening colonoscopy. As previously demonstrated, pre-selection by a gateopener FIT would go along with a significantly higher diagnostic yield and CRC risk reductions at the same number of conducted colonoscopies versus conventional screening colonoscopy.[9]

**Figure 1.**
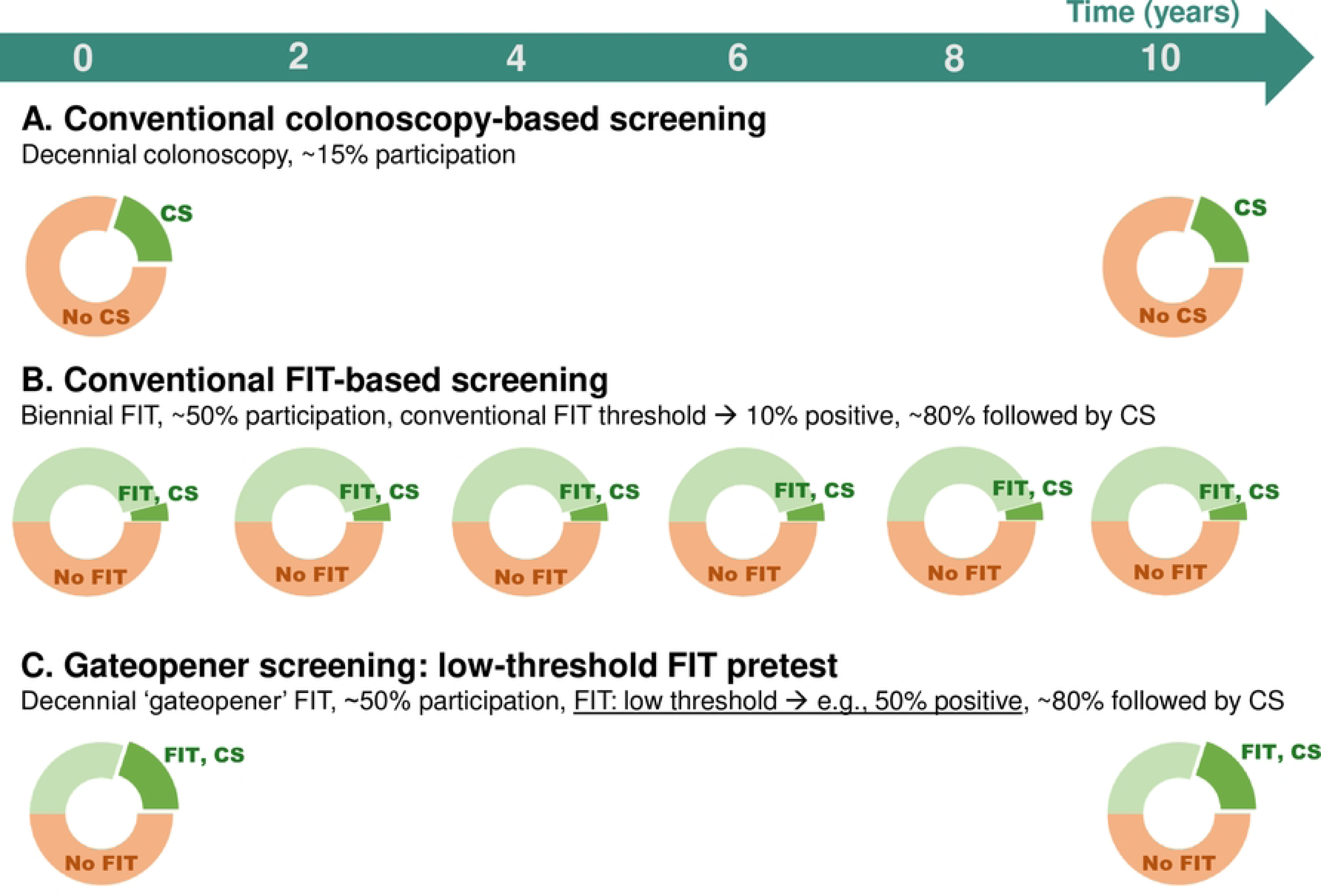
Numerical example of use of gateopener screening (C) as compared to conventional screening strategies (A, screening colonoscopy, B, biennial FIT) In the gateopener approach, instead of directly undergoing screening colonoscopy every 10 years, individuals would be invited to use a non-invasive, low-threshold pre-test, such as a FIT with very low hemoglobin detection thresholds yielding higher positivities (e.g., cut-off of 3-4 µg Hb/g, corresponding positivity: 50%, proposed in reference [9]). Only those above the threshold would be invited to follow-up with screening colonoscopy. CS: colonoscopy. FIT: fecal immunochemical test.

However, best possible use of the gateopener approach and its performance with respect to reduction of CRC incidence and mortality and most efficient use of colonoscopy resources remain to be explored. For example, our initial proposal focused on the use of a gateopener FIT with hemoglobin cut-offs of 3-4 µg/g feces yielding 50% positive test results.[9] However, depending on available colonoscopy capacities, trained staff and other resource constraints, higher hemoglobin cut-offs, which might nevertheless still maintain reasonably high sensitivity for detecting advanced CRC precursor lesions, might be better suited for some health care systems, also to reduce the proportion of false-positive gateopener FITs. Here, we sought to explore and compare the potential of various variants of gateopener screening for reducing CRC incidence and mortality and maximizing efficient use of colonoscopy resources.

## Materials and Methods

### Colorectal Cancer Multistate Simulation Model

To evaluate the implications of a gateopener screening approach at varying hemoglobin cut-offs, we used the <u>Co</u>lorectal Cancer Multistate <u>Si</u>mulation <u>Mo</u>del (COSIMO), a validated, Markov-based simulation tool,[10] in a hypothetical German population. Briefly, COSIMO simulates the natural history of CRC based on the process of precursor lesions developing into preclinical and then clinical cancer for a predefined number of years. Screening can interfere with the natural history of CRC (**Supplementary Figure 1**). A comprehensive documentation on the model’s structure, data sources and parameters is given in **Supplementary Appendix 1 and Supplementary Tables 1-3**. The model source code is available from our website.[11]

### Diagnostic Performance Parameters

We first derived sensitivity (the proportion of detected cases among all subjects with any adenomas or cancer) and specificity (the proportion of all subjects without adenomas or cancer correctly classified as such) parameters of SENTiFIT-FOB Gold (Sentinel Diagnostics, Milan, Italy), a commonly used FIT test, at hemoglobin (Hb) cut-offs of 10, 8, 6, 4, and 3 µg Hb per gram feces, respectively, which would correspond to positivity levels of approximately 20-50%. For reference, we also derived sensitivity and specificity at the manufacturer recommended cut-off of 17 µg/g. Cut-offs of 3-4 µg/g, corresponding to approximately 50% positivity, were originally proposed for use in the gateopener approach.[9]

Analyses were conducted based on 7,398 participants of screening colonoscopy in Germany recruited in the BLITZ study from 2008 through 2020 who had provided pre-colonoscopy stool samples for quantitative FITs as previously described.[7] Across all analyses, advanced adenomas were defined as adenomas with at least 1 of the following features: ≥ 1 cm in size, tubulovillous or villous components, or high-grade dysplasia. The BLITZ study was approved by the ethics committee of the Heidelberg medical faculty of Heidelberg University and the responsible ethics committees of the state medical boards. Written informed consent was obtained from each participant.

### Simulations

A range of scenarios was simulated to characterize the relative effectiveness of gateopener screening at varying cut-offs. In these scenarios, simulated cohorts of 100,000 previously unscreened women and men aged 50, 60, or 70 were assumed to be invited to undergo either:

1. conventional screening colonoscopy,
2. conventional biennial FIT screening,
3. gateopener screening at varying hemoglobin cut-offs (17, 10, 8, 6, 4, and 3 µg/g).

Sex-specific baseline neoplasm prevalences for each age of screening were extracted from a previous analysis of more than 4.4 million screening colonoscopies in the German screening-eligible population.[12] We assumed levels of uptake reflective for those reported for the German screening-eligible population, i.e., 15% uptake for conventional screening colonoscopy (offered in 10-year intervals) and 50% uptake for conventional biennial FIT and gateopener screening.[13] For biennial FIT screening, a 40% drop-out rate over time was assumed to account for the sporadic nature of adherence to FIT screening,[14] in line with reports in the literature.[15] Assuming 50% uptake over a 10-year period with 40% drop-out results in an approximate biennial uptake rate of 20% as reported for the German screen-eligible population.[13] We assumed that 80% of those with positive FIT test would make use of the follow-up colonoscopy offer, which reflects the observed compliance to diagnostic colonoscopy in real-world screening.[16]

### Outcomes

For each scenario, we assessed the number of prevented cases and deaths as compared to no screening after 10 years of follow-up, the numbers of required colonoscopies and FITs, as well as the numbers of required colonoscopies and FITs per prevented cases or death. We then calculated and compared the corresponding relative differences for gateopener screening scenarios to those for conventional screening scenarios (screening colonoscopy or biennial FIT).

### Patient and Public Involvement

Patients and the public were neither involved in the design and conduct of this study, nor in writing or editing of this document. Research at the German Cancer Research Center (DKFZ) is generally informed by a Patient Advisory Committee.

## Results

### Hemoglobin cut-offs

Sensitivities for any neoplasm (ANN, any adenoma or cancer), any advanced neoplasm (ADN, any advanced adenoma or cancer) and cancer as well as specificities for no finding at varying hemoglobin cut-offs are shown in **Table 1**. Sensitivities for ADN (as well as positivities) increased gradually with decreasing hemoglobin cut-offs (**Figure 2**) while specificities decreased first gradually (between cut-offs at 17 and 8 µg/g) and then rather steeply (between cut-offs at 8 and 3 µg/g).

**Table 1.** Diagnostic characteristics of SENTiFIT FOB Gold at varying hemoglobin cut-offs.

| Hemoglobin<br>cut-off<br>($\mu\text{g Hb/g}$) | Positivity<br>(%) | Sensitivity<br>(%) | | | Specificity<br>(%) |
| --- | --- | --- | --- | --- | --- |
|  |  | any<br>neoplasm | any advanced<br>neoplasm | preclinical<br>cancer | no<br>neoplasm |
| 17 | 10 | 19.6 | 39.4 | 90.7 | 93.3 |
| 10 | 15 | 27.4 | 48.5 | 96.3 | 88.9 |
| 8 | 19 | 32.1 | 53.4 | 98.1 | 85.1 |
| 6 | 29 | 40.8 | 59.5 | 98.1 | 74.7 |
| 4 | 44 | 54.5 | 68.3 | 98.1 | 58.2 |
| 3 | 51 | 60.4 | 72.7 | 98.1 | 51.1 |

Analyses based on 7,398 participants of screening colonoscopy in Germany from 2008 through 2020. advanced adenomas were defined as adenomas with at least 1 of the following features: $\geq 1$ cm in size, tubulovillous or villous components, or high-grade dysplasia.

**Figure 2.**
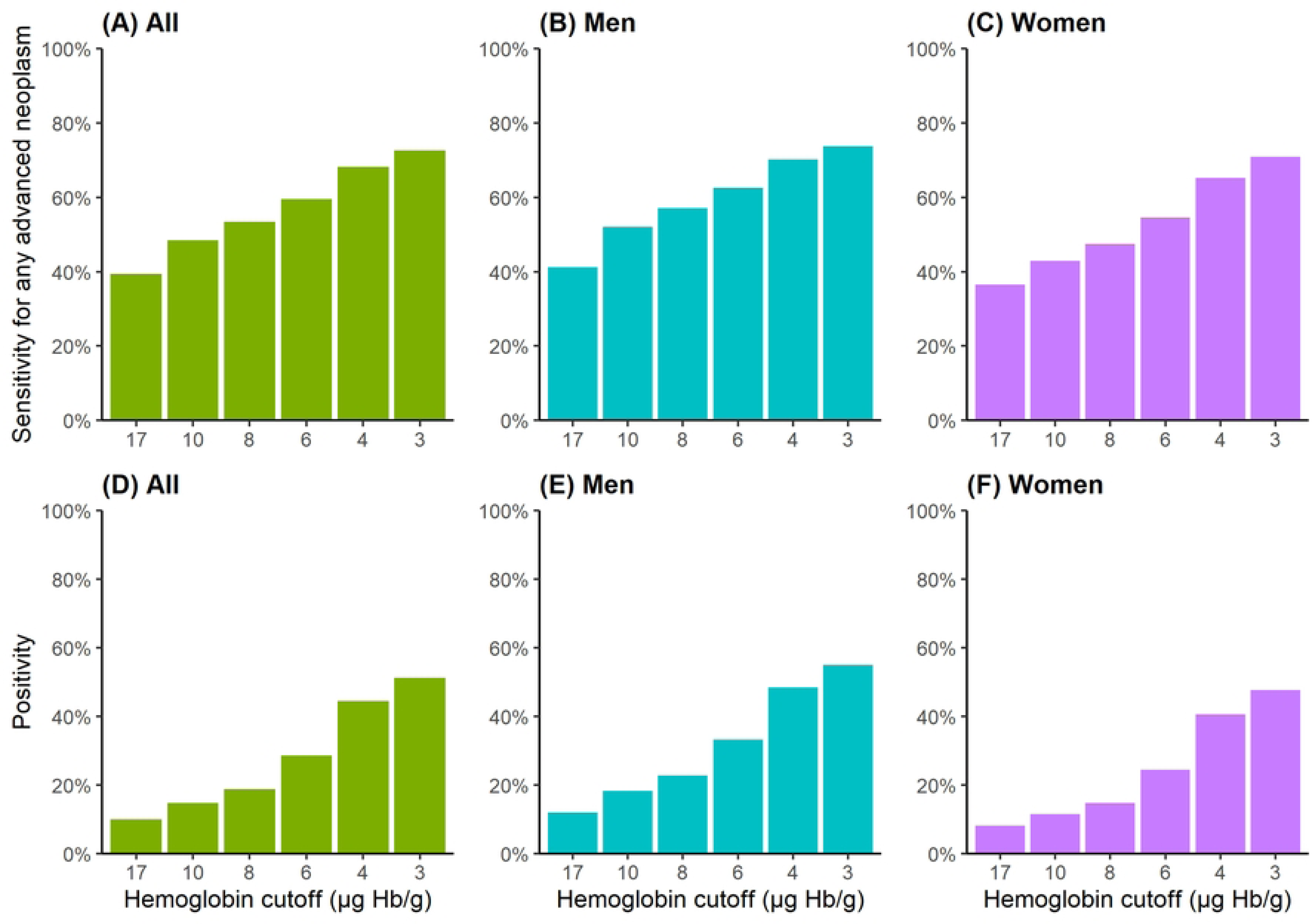
Sensitivity for any advanced neoplasm (advanced adenoma or cancer, panels A-C) and corresponding positivities (panels D-F) at varying FIT positivities.

### Comparison to conventional screening colonoscopy

Gateopener screening using a gateopener FIT with a cut-off of 6, 4 or 3 µg/g outperformed screening colonoscopy in terms of CRC incidence reduction, with 16-25%, 50-57% and 66-72% more prevented cases, respectively, across all ages (**Figure 3A, Supplementary Table 4**). Gateopener screening using a cut-off of 17 or 10 µg/g resulted in fewer and using a cut-off of 8 µg/g resulted in approximately equal numbers of prevented cases but, along with the other assessed hemoglobin thresholds, in significantly more prevented CRC deaths, with additional reductions reaching from +24% for a cut-off of 17 µg/g up to more than twice (+109%) the number of prevented deaths for conventional screening for a cut-off of 3 µg/g (**Figure 3B**). Screening using cut-offs of 17, 10, 8 or 6 µg/g also required fewer total colonoscopies (-6 to -66%). Absolute numbers of prevented cases or death by gateopener screening were less marked in women versus men, but relative gains were largely consistent across sexes (**Supplementary Figures 2 and 3**).

**Figure 3.**
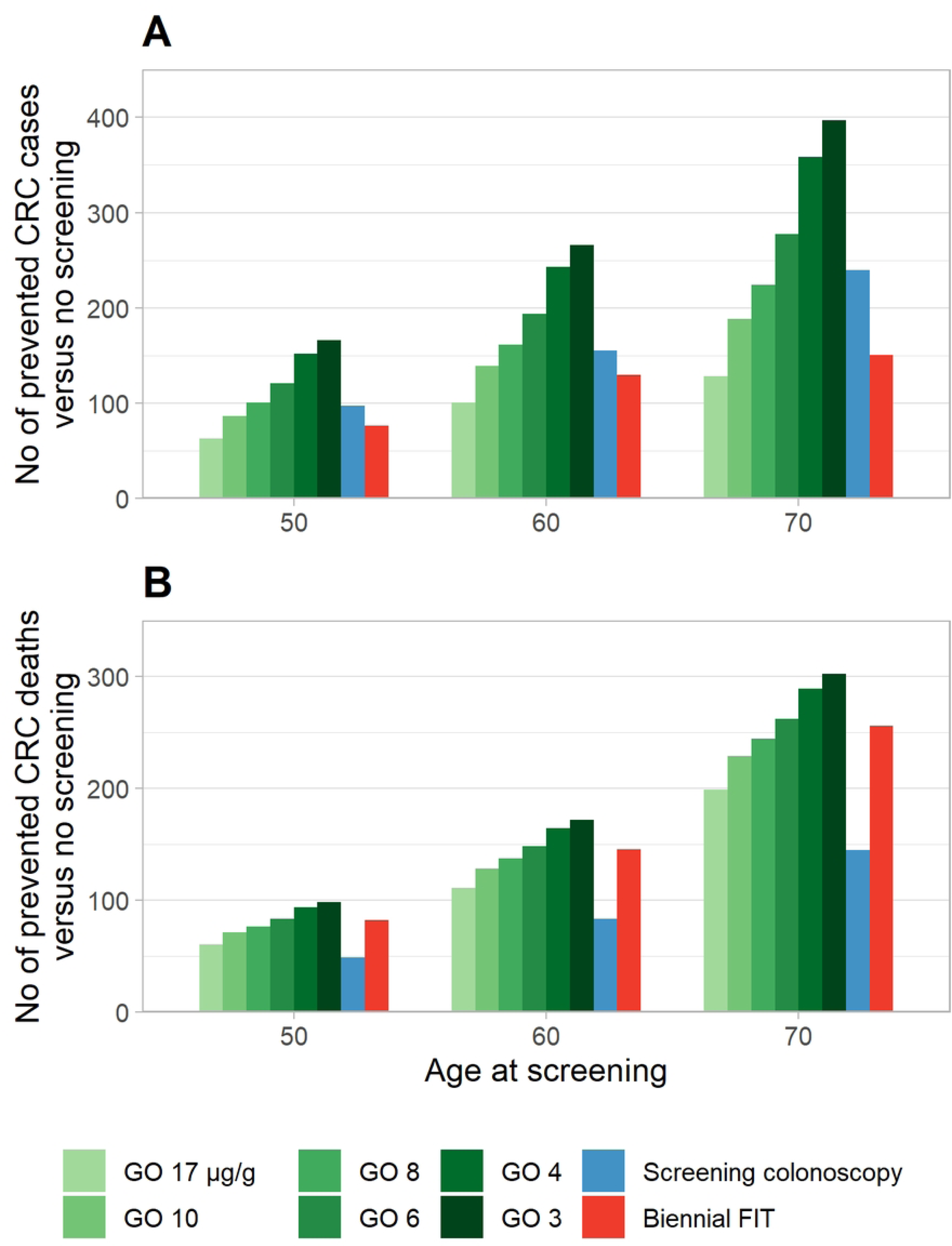
Numbers of prevented CRC cases and CRC deaths as compared to no screening at varying hemoglobin cut-offs, stratified by age. CRC, colorectal cancer; FIT, fecal immunochemical testing; GO, gateopener approach

### Comparison to conventional biennial FIT screening

Compared to biennial FIT screening, gateopener screening at a cut-off of 17 µg/g prevented 15-23% fewer CRC cases. At all other assessed cut-offs, gateopener screening prevented more CRC cases, with relative gains ranging from 7-163% across all assessed ages, with lower cut-offs associated with higher relative gains (**Figure 3A, Supplementary Table 4)**. Gateopener screening with cut-offs of 17 and 10 µg/g resulted in fewer (-11 to -26%), with cut-offs of 8 and 6 µg/g in approximately equal, and with cut-offs of 4 and 3 µg/g in significantly more (+12 to +21%) prevented deaths as compared to conventional biennial FIT screening, respectively (**Figure 3B**). Gateopener screening using a cut-off of 17 or 10 µg/g required 23-52% fewer total colonoscopies, while cut-offs of 6, 4 or 3 µg/g required 25-26%, 79-84%, and 102-110% more colonoscopies, respectively. At a cut-off of 8 µg/g, numbers of required colonoscopies were approximately comparable. The number of required FIT tests was more than halved (reduction by 54-57%) as compared to biennial FIT screening. Results were overall consistent across men and women and assessed ages.

### Numbers of colonoscopies per prevented case/per prevented death

Across all analyses, gateopener screening with cut-offs at 17, 10, 8 and 6 µg/g was consistently and partly substantially more efficient as compared to conventional screening strategies in terms of required numbers of colonoscopies per prevented CRC case (**Figure 4A**). Gateopener screening using a cut-off of 4 or 3 µg/g was consistently more efficient than screening colonoscopy for both sexes and more efficient than conventional biennial FIT screening in men. In women, higher efficiency versus biennial FIT only manifested at higher age (**Supplementary Figures 4 and 5**).

**Figure 4.**
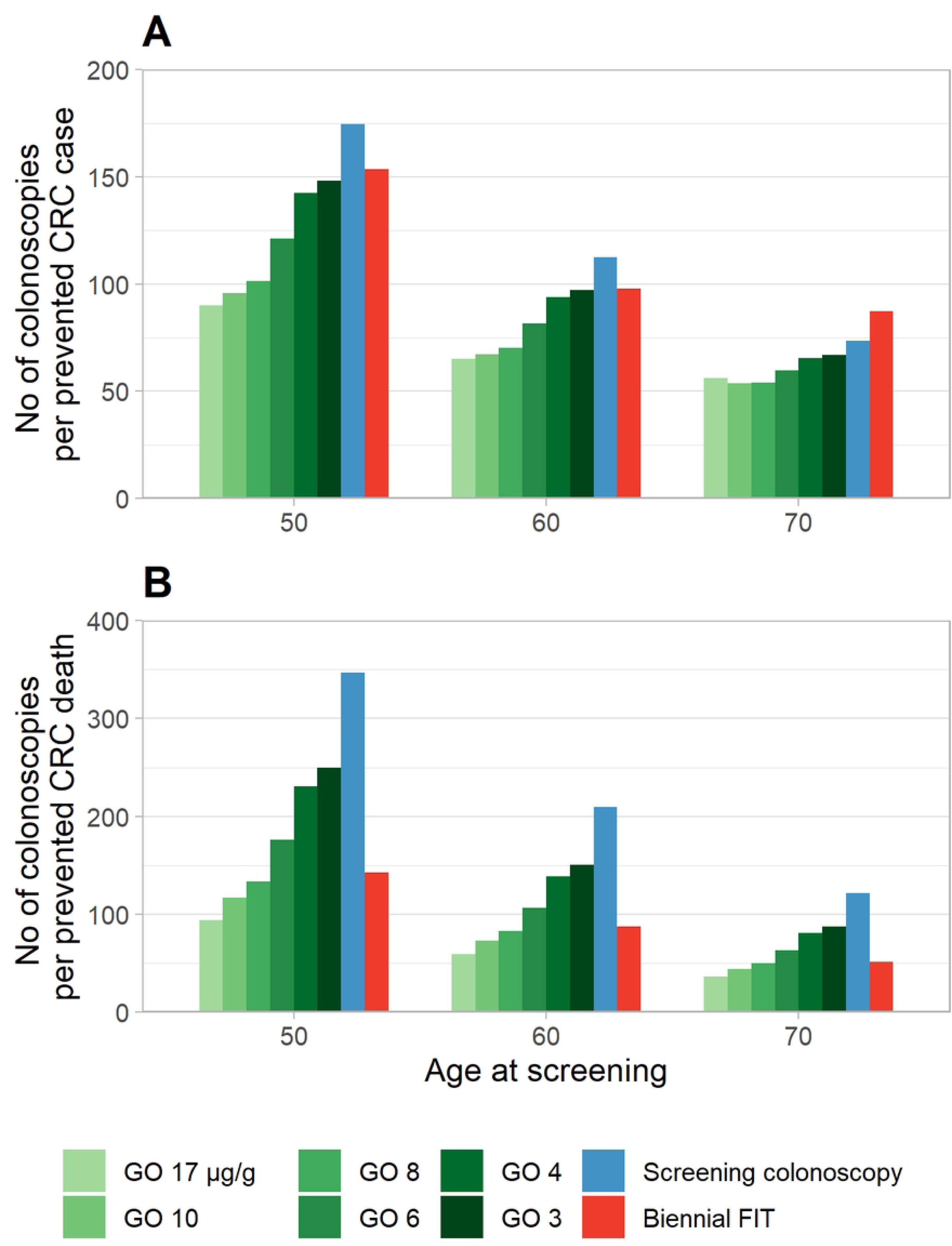
Numbers of colonoscopies per prevented CRC case and CRC death at varying hemoglobin cut-offs, stratified by age. CRC, colorectal cancer; FIT, fecal immunochemical testing; GO, gateopener approach

Gateopener screening with cut-offs at 17, 10, or 8 µg/g required considerably fewer colonoscopies per prevented CRC death as compared to conventional screening colonoscopy, and fewer to approximately equal numbers as compared to conventional biennial FIT testing (**Figure 4B**). Gateopener screening with cut-offs at 6, 4 or 3 µg/g was still consistently more efficient than screening colonoscopy but required more colonoscopies per prevented death than biennial FITs. For both measures (numbers per prevented case/deaths), differences across varying gateopener cut-offs tended to be become smaller with increasing age at screening.

## Discussion

Gateopener screening represents an innovative concept involving low barrier, low threshold non-invasive pretesting followed-up by screening colonoscopy for those with positive pre-test results. Gateopener pretesting with a single FIT, as proposed in this study, differs from conventional biennial FIT screening in two major aspects: first, it would be offered only in 10-year intervals, in line with recommendations on screening colonoscopy, the underlying main screening modality. Second, the hemoglobin threshold would be much lower than the one used for conventional FITs, implying significantly larger shares of positive (pre-)test results, with the specific threshold to be used depending on the specific healthcare systems circumstances, needs and resources. This study illustrates the strong advantages of gateopener screening for a broad range of thresholds: compared to conventional approaches, gateopener screening either would lead to higher proportions of prevented CRC cases or deaths, a more effective use of colonoscopies, or, for a range of hemoglobin cut-offs, even both.

### Findings in Context

Attributable to demographic aging, unfavorable trends in major risk factors such as Western dietary habits (such as high fat and sugar, low fiber content; red meat consumption; highly processed food),[17] as well as the likely existence of birth cohort effects implying increasing incidence in younger generations,[18,19] the number of worldwide CRC cases and deaths is set to rise significantly in the future.[20–22] At the same time, an increasing number of healthcare systems faces strong constraints related to budget, workforce, and capacities for cancer care,[23,24] exacerbated by competing priorities and ever rising costs to treat CRC.[25,26]

These alarming developments point to an urgent need to rethink current approaches to CRC screening. In principle, the tools to mitigate if not counteract the predicted CRC epidemic already exist. Full adherence to either of the most popular conventional options, screening endoscopy or biennial stool-test-based screening, might reduce lifelong CRC risks by 70-80%.[27,28] However, screening effectiveness and successful screening implementation in practice is key, and both conventionally followed approaches, colonoscopy-based and FIT-based screening, come along with limitations in this respect.

Although screening colonoscopy is the gold standard examination for detecting CRC and its precursor lesions, it is a relative burdensome and invasive procedure, whose uptake rates are low, even in countries where it is offered free of charge as primary screening exam, such as Germany, where 10-year use rates of 10-20% have been observed.[13] Few countries, including the US, report considerably higher endoscopy use rates,[4] but still approximately 60% of CRC deaths in the US are attributable to the non-use of screening.[29,30] In addition, only few individuals benefit from this in relative terms resource-intensive procedure, making it rather inefficient on a population level. According to previous estimates, approximately 200 and 400 screen-eligible men and women aged 50 years, respectively, would need to be screened to detect a single case of cancer.[9] In Germany, costs for a colonoscopy are approximately 30-times the costs for a fecal test (but still cost-saving).[31,32]

Conversely, uptake of fecal-test-based screening, e.g., when offering FITs, is higher, owing to its non-invasive nature and more convenient conduct. However, conventional FIT screening has only limited ability to reduce CRC incidence, as, when used at manufacturer recommended hemoglobin thresholds, sensitivity is high for preclinical cancers but suboptimal for precursor lesions.[7,8] FIT screening therefore needs to be repeated annually or every other year, requiring substantial resources and impairing its practicability over a life-long timeframe. In effect, a considerable proportion of users follows suboptimal sporadic uptake patterns, and adherence is highest in the first screening round.[14,33,34]

These limitations imply that conventional screening approaches alone are unlikely to mount a sufficient response to the predicted strong increase of the future CRC burden. Even in a country such as Germany with relatively high saturation and capacities, by one estimate, screening colonoscopy uptake would need to increase to more than 3-times current levels to avoid increasing case numbers.[18] Such a large increase is unrealistic, though somewhat higher uptake levels could plausibly be achieved, e.g., by implementing a well-designed organized screening program.[35] Yet, real progress will need to be brought about by redesigning CRC screening involving innovative approaches leading to higher uptake and greater effectiveness.

Gateopener screening is a strong candidate to address this gap. It unites the advantages of FIT-based screening, i.e., high uptake and low barriers of use (as only a single FIT is needed every ten years, which should simplify personal invitations and allow for postal provision, which could lead to even higher uptake[6]), with those of primary screening colonoscopy, i.e., detection of precursor lesions with high sensitivity. As demonstrated in our analyses, another key feature of gateopener screening is the ability to direct colonoscopy capacities to those most likely to benefit while maintaining or likely improving overall screening outcomes.

In our study, by making use of the (typically neglected) quantitative potential of modern FITs, we illustrated several possibilities to implement gateopener screening. From a public health perspective, the chosen hemoglobin cut-off for the gateopener FITs reflects a continuum: on the end with low hemoglobin thresholds, screening outcomes, i.e., numbers of prevented cases and deaths are maximized, at the cost of somewhat lower screening efficiency (higher share of false-positive tests and higher numbers needed to screen). At the other end, with high hemoglobin thresholds, screening efficiency is highest (lower numbers needed to screen), at the cost of fewer overall prevented cases and deaths. It should be noted that in virtually all simulations, regardless of the eventually chosen cut-off, gateopener screening outperformed conventional screening on either or both metrics.

We deliberately abstained from identifying an ‘optimal’ hemoglobin cut-off. A strength the gateopener approach is that each healthcare system can implement it according to individual priorities, preferences and resources. For instance, relatively resource- and capacity-strong systems, such as the US or Germany, where primary screening colonoscopy is already offered, might opt for rather low hemoglobin thresholds. This would allow to further improve screening outcomes (i.e., further reduce CRC incidence and mortality) while maintaining adequately high levels of screening efficiency. As capacities become tighter due to anticipated effects of demographic aging, this approach would allow for gradual adjustments of thresholds to reflect available capacities. Also, gateopener screening could be an excellent alternative to annual or biennial FIT screening, as offered across many European screening programs. For some cut-offs, screening outcomes and numbers of required colonoscopies were approximately comparable to biennial FIT screening, but the number of required FITs was more than halved, making gateopener screening the far more efficient (and also more convenient) option. Finally, gateopener screening starting at moderate or high hemoglobin cut-offs could be an excellent choice for countries with yet pending or uncomplete roll-out of CRC screening offers. The ability to adjust for cut-offs represents a strong instrument until rollout is implemented or capacities are fully built up.

It should be noted that the comparably high false-positive rate of low cut-off gateopener screening should not be used as argument against this approach. For primary screening colonoscopy, the established gold standard, approximately 70-80% of individuals undergo the procedure without any finding, and close to 90% have no advanced finding.[36] More than 2000 and 3500 colonoscopies in 50-year-old men and women, respectively, were estimated to be needed to detect a single case of cancer in those who would hypothetically undergo screening colonoscopy but were prevented from doing so due to negative pretesting with a gateopener FIT using very low cut-offs of 3-4 µg/g.[9] In other words, most individuals do not benefit from conventional screening colonoscopies, and gateopener screening will significantly improve screening efficiency. However, clear patient communication on the advantages and specifics of gateopener screening will be key, ideally in a well-formulated invitation letter. Smart program design involving such inviting letters as well as postal provision of FITs would likely substantially improve the uptake and acceptance.[6]

### Limitations

Specific limitations of COSIMO have been described previously.[9,10] Briefly, major limitations concern simplifying model assumptions and uncertainties related to input parameters. For instance, true adenoma miss rates at colonoscopy are unknown, and we used the best available evidence from a systematic review and meta-analysis for approximation.[1] Furthermore, as subsite specific data was not available in the registry data used to inform COSIMO model parameters,[10] no stratified analysis for distal versus proximal CRC was possible, which would have allowed for comparisons to screening sigmoidoscopy, another broadly established endoscopic screening intervention.

In addition, simulations were conducted for a screen-eligible population in Germany (for which the simulation model COSIMO was previously calibrated and validated[10]), and baseline prevalences as well as adenoma and cancer incidence rates might vary for other populations with varying distributions of CRC risk factors. On a similar note, input adherence patterns were derived from German data sources, and uptake will likely vary across countries and healthcare systems. However, in our study, key outcome parameters are reported on a relative effect scale and should be interpreted relative to the measured effectiveness of conventional screening approaches, and it appears plausible that the relative effectiveness of gateopener screening will be consistent across populations. A pilot study would be needed to assess the plausibility of uptake assumptions as well as the overall feasibility of the gateopener concept. Further research should also consider health economic considerations, including the cost increase associated with a potential postal delivery of gateopener FIT test kits, as well as likely cost savings due to improved CRC prevention and early diagnosis.

### Conclusion

Gateopener screening is an innovative concept for prevention and early detection of CRC. The approach involves low barrier, low threshold non-invasive pretesting using a single ‘gateopener’ FIT followed-up by screening colonoscopy for those with positive pre-test results. This ‘gateopener FIT’ may be adjusted by modifying hemoglobin cut-offs indicating when such pre-test will be considered positive, whereby relatively lower cut-offs will imply higher numbers of prevented CRC cases and deaths, and relatively higher cut-offs will imply higher screening effectiveness (i.e., fewer colonoscopies to detect one individual with an advanced finding). This study illustrates that gateopener screening at varying cut-offs will outperform conventional screening approaches by offering considerably stronger reduction of CRC incidence and mortality rates as well as considerably increased screening effectiveness. The feasibility of the concept should be assessed by a pilot study in real-life practice.

## Declarations

### Ethics approval

The BLITZ study was approved by the ethics committees of Heidelberg University (178/2005) and the state medical chambers of Baden-Württemberg (M118-05-f), Saarland (217/13), Rhineland Palatinate (837.047.06(5145)) and Hesse (MC 254/2007).

### Consent to participate and for publication

All participants in BLITZ provided written informed consent.

### Availability of data, code and material

All analyses relevant to the study are included in the article or uploaded as supplementary information. The model source code is freely available from the DKFZ website (https://www.dkfz.de/de/klinepi/download/index.html).

### Authors’ contributions

HB and TH designed the study and developed the methodology. TH conducted the statistical analyses and drafted the manuscript. All authors critically reviewed the manuscript, contributed to its revision, and approved the final version submitted. The researchers are independent from funders. All authors had full access to all of the data (including statistical reports and tables) used for the study and can take responsibility for the integrity of the data and the accuracy of the data analysis.

## References

1. Zhao S, Wang S, Pan P, Xia T, Chang X, Yang X, et al. Magnitude, Risk Factors, and Factors Associated With Adenoma Miss Rate of Tandem Colonoscopy: A Systematic Review and Meta-analysis. Gastroenterology. 2019;156: 1661–1674.e11. doi:10.1053/j.gastro.2019.01.260

2. Lin JS, Perdue LA, Henrikson NB, Bean SI, Blasi PR. Screening for Colorectal Cancer: Updated Evidence Report and Systematic Review for the US Preventive Services Task Force. J Am Med Assoc. 2021;325: 1978–1998. doi:10.1001/jama.2021.4417

3. Imperiale TF, Abhyankar PR, Stump TE, Emmett TW. Prevalence of Advanced, Precancerous Colorectal Neoplasms in Black and White Populations: A Systematic Review and Meta-analysis. Gastroenterology. 2018;155: 1776–1786.e1. doi:10.1053/j.gastro.2018.08.020

4. Cardoso R, Niedermaier T, Chen C, Hoffmeister M, Brenner H. Colonoscopy and Sigmoidoscopy Use among the Average-Risk Population for Colorectal Cancer: A Systematic Review and Trend Analysis. Cancer Prev Res (Phila). 2019;12: 617–630. doi:10.1158/1940-6207.CAPR-19-0202

5. Cardoso R, Guo F, Heisser T, Hoffmeister M, Brenner H. Utilisation of Colorectal Cancer Screening Tests in European Countries by Type of Screening Offer: Results from the European Health Interview Survey. Cancers (Basel). 2020;12: E1409. doi:10.3390/cancers12061409

6. Gruner LF, Amitay EL, Heisser T, Guo F, Niedermaier T, Gies A, et al. The Effects of Different Invitation Schemes on the Use of Fecal Occult Blood Tests for Colorectal Cancer Screening: Systematic Review of Randomized Controlled Trials. Cancers (Basel). 2021;13: 1520. doi:10.3390/cancers13071520

7. Gies A, Cuk K, Schrotz-King P, Brenner H. Direct Comparison of Diagnostic Performance of 9 Quantitative Fecal Immunochemical Tests for Colorectal Cancer Screening. Gastroenterology. 2018;154: 93–104. doi:10.1053/j.gastro.2017.09.018

8. Brenner H, Qian J, Werner S. Variation of diagnostic performance of fecal immunochemical testing for hemoglobin by sex and age: results from a large screening cohort. Clin Epidemiol. 2018;10: 381–389. doi:10.2147/CLEP.S155548

9. Heisser T, Cardoso R, Niedermaier T, Hoffmeister M, Brenner H. Making colonoscopy-based screening more efficient: A “gateopener” approach. International Journal of Cancer. 2023;152: 952–961. doi:10.1002/ijc.34317

10. Heisser T, Hoffmeister M, Brenner H. Effects of screening for colorectal cancer: Development, documentation and validation of a multistate Markov model. Int J Cancer. 2021;148: 1973–1981. doi:10.1002/ijc.33437

11. German Cancer Research Center (DKFZ), Department Clinical Epidemiology and Aging Reserach. Download Page for COSIMO Source Code. Jun 2023 [cited 20 Jun 2023]. Available: https://www.dkfz.de/en/klinepi/download/index.html

12. Brenner H, Altenhofen L, Stock C, Hoffmeister M. Prevention, early detection, and overdiagnosis of colorectal cancer within 10 years of screening colonoscopy in Germany. Clin Gastroenterol Hepatol. 2015;13: 717–23. doi:10.1016/j.cgh.2014.08.036

13. Kretschmann J, El Mahi C, Lichtner F, Hagen B. Früherkennungskoloskopie. Jahresbericht 2019. [Screening Colonoscopy. Annual Report 2019]. 2 Dec 2021 [cited 20 Jun 2023]. Available: https://www.zi.de/publikationen/studien/darmkrebs-frueherkennung

14. Heisser T, Cardoso R, Guo F, Moellers T, Hoffmeister M, Brenner H. Strongly Divergent Impact of Adherence Patterns on Efficacy of Colorectal Cancer Screening: The Need to Refine Adherence Statistics. Clin Transl Gastroenterol. 2021;12: e00399. doi:10.14309/ctg.0000000000000399

15. Murphy CC, Sen A, Watson B, Gupta S, Mayo H, Singal AG. A Systematic Review of Repeat Fecal Occult Blood Tests for Colorectal Cancer Screening. Cancer Epidemiol Biomarkers Prev. 2020;29: 278–287. doi:10.1158/1055-9965.EPI-19-0775

16. Gingold-Belfer R, Leibovitzh H, Boltin D, Issa N, Tsadok Perets T, Dickman R, et al. The compliance rate for the second diagnostic evaluation after a positive fecal occult blood test: A systematic review and meta-analysis. United European Gastroenterol J. 2019;7: 424–448. doi:10.1177/2050640619828185

17. Carr PR, Weigl K, Edelmann D, Jansen L, Chang-Claude J, Brenner H, et al. Estimation of Absolute Risk of Colorectal Cancer Based on Healthy Lifestyle, Genetic Risk, and Colonoscopy Status in a Population-Based Study. Gastroenterology. 2020;159: 129–138.e9. doi:10.1053/j.gastro.2020.03.016

18. Heisser T, Hoffmeister M, Tillmanns H, Brenner H. Impact of demographic changes and screening colonoscopy on long-term projection of incident colorectal cancer cases in Germany: A modelling study. The Lancet Regional Health – Europe. 2022;20: 100451. doi:10.1016/j.lanepe.2022.100451

19. Siegel RL, Torre LA, Soerjomataram I, Hayes RB, Bray F, Weber TK, et al. Global patterns and trends in colorectal cancer incidence in young adults. Gut. 2019;68: 2179– 2185. doi:10.1136/gutjnl-2019-319511

20. Arnold M, Sierra MS, Laversanne M, Soerjomataram I, Jemal A, Bray F. Global patterns and trends in colorectal cancer incidence and mortality. Gut. 2017;66: 683–691. doi:10.1136/gutjnl-2015-310912

21. Ferlay J, Laversanne M, Ervik M, Lam F, Colombet M, Mery L, et al. Global Cancer Observatory: Cancer Tomorrow. 2020. Available: https://gco.iarc.fr/tomorrow

22. Sung H, Ferlay J, Siegel RL, Laversanne M, Soerjomataram I, Jemal A, et al. Global Cancer Statistics 2020: GLOBOCAN Estimates of Incidence and Mortality Worldwide for 36 Cancers in 185 Countries. CA Cancer J Clin. 2021;71: 209–249. doi:10.3322/caac.21660

23. Kanavos P, Schurer W. The dynamics of colorectal cancer management in 17 countries. Eur J Health Econ. 2010;10: 115–129. doi:10.1007/s10198-009-0201-2

24. Henderson RH, French D, Maughan T, Adams R, Allemani C, Minicozzi P, et al. The economic burden of colorectal cancer across Europe: a population-based cost-of-illness study. Lancet Gastroenterol Hepatol. 2021;6: 709–722. doi:10.1016/S2468-1253(21)00147-3

25. Ladabaum U, Mannalithara A, Meester RGS, Gupta S, Schoen RE. Cost-Effectiveness and National Effects of Initiating Colorectal Cancer Screening for Average-Risk Persons at Age 45 Years Instead of 50 Years. Gastroenterology. 2019;157: 137–148. doi:10.1053/j.gastro.2019.03.023

26. Hofmarcher T, Lindgren P, Wilking N, Jönsson B. The cost of cancer in Europe 2018. European Journal of Cancer. 2020;129: 41–49. doi:10.1016/j.ejca.2020.01.011

27. Ladabaum U, Dominitz JA, Kahi C, Schoen RE. Strategies for Colorectal Cancer Screening. Gastroenterology. 2020;158: 418–432. doi:10.1053/j.gastro.2019.06.043

28. Heisser T, Hoffmeister M, Brenner H. Model based evaluation of long-term efficacy of existing and alternative colorectal cancer screening offers: A case study for Germany. Int J Cancer. 2022;150: 1471–1480. doi:10.1002/ijc.33894

29. Meester RGS, Doubeni CA, Lansdorp-Vogelaar I, Goede SL, Levin TR, Quinn VP, et al. Colorectal cancer deaths attributable to nonuse of screening in the United States. Ann Epidemiol. 2015;25: 208–213.e1. doi:10.1016/j.annepidem.2014.11.011

30. Siegel RL, Wagle NS, Cercek A, Smith RA, Jemal A. Colorectal cancer statistics, 2023. CA: A Cancer Journal for Clinicians. 2023;73: 233–254. doi:10.3322/caac.21772

31. Ran T, Cheng CY, Misselwitz B, Brenner H, Ubels J, Schlander M. Cost-Effectiveness of Colorectal Cancer Screening Strategies-A Systematic Review. Clin Gastroenterol Hepatol. 2019;17: 1969–1981.e15. doi:10.1016/j.cgh.2019.01.014

32. Heisser T, Simon A, Hapfelmeier J, Hoffmeister M, Brenner H. Treatment Costs of Colorectal Cancer by Sex and Age: Population-Based Study on Health Insurance Data from Germany. Cancers. 2022;14: 3836. doi:10.3390/cancers14153836

33. Jensen CD, Corley DA, Quinn VP, Doubeni CA, Zauber AG, Lee JK, et al. Fecal Immunochemical Test Program Performance Over 4 Rounds of Annual Screening: A Retrospective Cohort Study. Ann Intern Med. 2016;164: 456–463. doi:10.7326/M15-0983

34. Zorzi M, Hassan C, Capodaglio G, Fedato C, Montaguti A, Turrin A, et al. Long-term performance of colorectal cancerscreening programmes based on the faecal immunochemical test. Gut. 2018;67: 2124–2130. doi:10.1136/gutjnl-2017-314753

35. Toes-Zoutendijk E, van Leerdam ME, Dekker E, van Hees F, Penning C, Nagtegaal I, et al. Real-Time Monitoring of Results During First Year of Dutch Colorectal Cancer Screening Program and Optimization by Altering Fecal Immunochemical Test Cut-Off Levels. Gastroenterology. 2017;152: 767–775.e2. doi:10.1053/j.gastro.2016.11.022

36. Heisser T, Kretschmann J, Hagen B, Niedermaier T, Hoffmeister M, Brenner H. Prevalence of Colorectal Neoplasia 10 or More Years After a Negative Screening Colonoscopy in 120 000 Repeated Screening Colonoscopies. JAMA Internal Medicine. 2023;183: 183–190. doi:10.1001/jamainternmed.2022.6215

